# Temporal trends in primary care-recorded self-harm during and beyond the first year of the COVID-19 pandemic: time series analysis of electronic healthcare records for 2.8 million patients in the Greater Manchester Care Record

**DOI:** 10.1101/2021.06.23.21259400

**Authors:** Sarah Steeg, Lana Bojanić, George Tilston, Richard Williams, David A. Jenkins, Matthew J. Carr, Niels Peek, Darren M. Ashcroft, Nav Kapur, Jennifer Voorhees, Roger T. Webb

## Abstract

**Background:** Surveillance of clinically treated self-harm episode frequency is an important component of suicide prevention in the dynamic context of COVID-19. Studies published to date have investigated the initial months following the onset of the pandemic, despite national and regional restrictions persisting to Summer 2021.

**Methods:** We conducted a descriptive time series analysis utilising data from the Greater Manchester Care Record, which contains de-identified, primary care health records of 2.8 million patients. Counts of incident and all episodes of self-harm recorded between 1^st^ January 2019 and 31^st^ May 2021 were made for all patients, with stratification by sex, age group, ethnicity, and index of multiple deprivation (IMD) quintile and examination of overall differences by national and regional restriction phases.

**Findings:** Between 1st January 2019 and 31^st^ May 2021, 33,444 episodes of self-harm by 13,148 individuals were recorded. Frequency ratios of incident and all episodes of self-harm were 0.59 (95% CI 0.51 to 0.69) and 0.69 (CI 0.63 to 0.75) respectively in April 2020 compared to February 2020. Between August 2020 and May 2021 frequency ratios were 0.92 (CI 0.88 to 0.96) for incident episodes and 0.86 (CI 0.84 to 0.88) for all episodes compared to the same months in 2019. Reductions were largest among men and people living in the most deprived neighbourhoods. An increase in all-episode self-harm (frequency ratio 1.09, CI 1.03 to 1.16) was observed for adolescents aged 10-17 between August 2020 and May 2021.

**Interpretation:** The COVID-19 pandemic has had a sustained impact on help seeking for self-harm. Reductions in primary care recorded self-harm have implications for clinicians’ ability to assess the needs and risks of individuals. Some patients may be experiencing prolonged untreated deterioration in their mental health while other groups are presenting in higher numbers. Our findings have important implications for primary care and mental health services in manging ongoing demand.

**Funding:** UKRI COVID-19 Rapid Response Initiative (grant reference COV0499), University of Manchester Presidential Fellowship (SS), and NIHR Greater Manchester Patient Safety Translational Research Centre.

## Introduction

Surveillance of temporal trends in the frequency of clinically treated self-harm episodes has been identified as an important component of suicide prevention, ^1,2^ particularly so during the rapidly evolving context of the COVID-19 pandemic. ^3^ Evidence indicates that people’s confidence in the capacity of health services to manage demand has fluctuated through the course of the pandemic. ^4^ Several studies have examined local impacts on help seeking for health conditions in the months subsequent to the first wave of COVID-19 in the UK. Marked reductions in frequency of common mental and physical health conditions being recorded in electronic healthcare records following the onset of the pandemic in March 2020 have been reported in Salford, Greater Manchester, North West of England. ^5^ Furthermore, local reductions in emergency department visits for mental illness and self-harm have been reported in several European countries. ^6-8^ An investigation conducted using the UK’s Clinical Practice Research Datalink (CPRD) found that absolute reductions in primary care-recorded self-harm episode counts were largest for general practices in the most deprived localities. ^9^ This study found that, across the UK as a whole, frequency of primary care-recorded self-harm episodes reduced sharply in April 2020, by around a third. By July 2020, counts had almost returned to pre-pandemic levels and remained so until August 2020. However, while these findings are broadly representative of the UK population, it is not known how help seeking was affected in UK regions experiencing more prolonged COVID-19 containment restrictions from summer 2020. Furthermore, these findings do not provide information about use of health services beyond autumn 2020, through to spring 2021.

In addition to altering the delivery of healthcare services and help-seeking behaviour, the COVID-19 pandemic has also affected incidence and prevalence of mental illness and self-harm in communities. Increases in mental distress in UK respondents were found in April 2020 compared to predictions based on prior trends. ^10^ While most people who had experienced worsening mental health early in the pandemic had improved by October 2020, around one in nine continued to experience mental distress. ^11^ However, this study’s observation period did not include the later months of the pandemic when the UK returned to national lockdowns. A report by the Office for National Statistics found that the prevalence of depressive symptoms was 19% higher between January and March 2021 compared to November 2020. ^12^

Data from 21 high- and upper-middle-income countries has indicated that there was no increase in the number of suicide deaths during the initial phase of the pandemic in spring 2020, ^13^ though an increase in the suicide rate in Japan was found during later months of summer 2020. ^14^ later study found an increase in Likewise, a ‘living’ systematic review investigating the impacts of COVID-19 on self-harm and suicide internationally has found little evidence of rises in suicide or self-harm incidence during the months of the COVID-19 crisis, though the authors note that better quality, peer-reviewed studies are required to strengthen the evidence base. ^15^ Although one study found changes in reported levels of suicidal ideation, ^16^ another found no overall change in the proportions of adults reporting that they had self-harmed, ^4^ though it did report higher levels among certain groups including younger people, those with a lower income and people with a diagnosed mental or physical illness. However, findings from self-report surveys do not provide any information about help-seeking behaviour or receipt of healthcare. This is important because people who have self-harmed have increased risks of suicide and they therefore require early intervention. ^17^ There are also few studies examining the later periods of the pandemic as it progressed through the 2020-2021 winter and 2021 spring.

It is particularly important to examine temporal trends in the frequency of recorded self-harm episodes at regional level throughout the COVID-19 epoch, beyond the first national lockdown period and the relatively relaxed summer months of 2020 during which the early mitigation restrictions were relaxed. Following the first set of COVID-19 restrictions in March, April and May 2020, regions of the UK experienced varying levels of containment measures. For example, several areas in the North West of England, including Greater Manchester, were subject to additional restrictions during July to September 2020. ^18^ It is not known how the second national lockdown in November 2020, the implementation of Tier 3 restrictions in Greater Manchester in December 2020 and the third national lockdown in January to March 2021 impacted help seeking for self-harm. We therefore aimed to examine temporal trends in monthly counts of primary care-recorded self-harm episodes from January 2019 to the end of May 2021 for all GP-registered residents in the socially diverse but relatively deprived conurbation of Greater Manchester in the North West of England. We examined differences in recorded frequencies of self-harm by broad phases of national and regional restrictions. We also examined differences in temporal trends according to sex, age group, ethnicity and Index of Multiple Deprivation (IMD) quintile.

## Methods

### Data sources, study design and data access approval

This descriptive time series analysis was conducted using the Greater Manchester Care Record (GMCR), ^19^ which includes information from primary care electronic healthcare records. The dataset holds information on approximately 2.8 million GP-registered patients. Research protocols must be approved and be in line with the national Control of Patient Information (COPI) notice, which gives NHS organisations a legal requirement to share data for the purposes of the COVID-19 response.

### Definitions, measurements and clinical coding

We included patient records from 1^st^ January 2019, the earliest date from when complete data were available, to 31^st^ May 2021, and we extracted monthly counts of relevant clinical codes for self-harm entered into patients’ records throughout that period (1^st^ January 2019 to 31^st^ May 2021). Data were extracted on June 14^th^ 2021 to capture episodes entered into the patient record up to two weeks after the date of self-harm and to account for the delay between entry in the patient record and appearing in the GMCR. Patient clinical information in the GMCR comes from a variety of GP systems and so is recorded using a combination of Read v2, CTV3, EMIS and SNOMED codes (for codes, see https://github.com/rw251/gm-idcr/tree/master/shared/clinical-code-sets/patient/selfharm-episodes/1). Some codes would have resulted from a hospital presentation that was subsequently entered into the patient’s primary care record. The prevalence of self-harm codes recorded across the three GP systems used in Greater Manchester (EMIS, TPP and Vision) were compared as a validation of the robustness of the codes used. The percentage of patients in the GMCR with a self-harm code in their GP record varied by less than ten percent across the three GP systems. Codes entered multiple times on the same day in a patient’s record were considered as a single episode. We summed incident (i.e. first-recorded) and all-episodes of self-harm entered in patients’ records, with the former identified using all previous entries in a patient’s primary care record, including those prior to 2019, as a look back window. A broad definition of self-harm was applied, capturing episodes of varying suicidal intent, in line with the definition used in UK National Institute for Health and Care Excellence (NICE) guidance. ^20^ The code lists used were verified by senior clinical academics in a previous study. ^9^

Age groups and ethnicity categories were collapsed to avoid reporting counts lower than 10. Frequencies of recorded incident self-harm episodes were stratified by sex, age group (adolescents: 10-17; young adults through early middle age: 18-45; middle aged and older adults: 46 years and older), index of multiple deprivation (IMD) quintile, and ethnicity (White vs. Black/Black British, Asian/Asian British, other and mixed ethnic group combined). The same categories were used for frequencies of all self-harm events with the exception of age group for which counts were sufficiently large to enable the 45+ years group to be split into 45-64 and 65 years and over. We focussed on two broad comparison periods: (i) February 2020 vs. April 2020, the month before the onset of the pandemic compared to the month where previous research has shown there was a large reduction in frequency of health services usage, and (ii) August 2020, from when service use had broadly returned to expected levels to May 2021 vs. the same months in 2019. ^9^ We also examined frequency ratios between different phases of national and regional restrictions and equivalent calendar month periods in 2019.

### Patient and public involvement

Four service users and carers with lived experience of mental health services worked with the research team to interpret the findings of this study. The group is linked with the National Institute Health Research Greater Manchester Patient Safety Translational Research Centre (NIHR GM PSTRC). The GRIPP2 S-F ^21^ checklist was used to report involvement.

### Data analyses

The data were structured as time-series data by calculating incident and all-episode self-harm frequencies, separately, per month. Subgroup analyses were conducted by stratifying the monthly aggregated data. Exploratory analyses were conducted to determine the age and ethnicity categories used in our study to ensure we did not report on groups lower than 10 patients, while aiming to apply informative and meaningful groupings. For analyses stratified by sex, age group, IMD quintile and ethnicity, episodes with missing data for these variables were excluded list-wise. Frequency ratios and their 95% confidence intervals were calculated using negative binomial regression. Data analysis was conducted in R version 3.6.3 and Stata SE v15.1. Authors LB, SS and GT had access to the data used for the study and GT had access to the database used to create the study population. We followed RECORD (REporting of studies Conducted using Observational Routinely-collected health Data) guidance (see online supplement). ^22^ The GMCR Research Governance Group approved the protocol (reference RQ-029) for this study in March 2021. All patient data were de-identified and informed consent was therefore not needed.

## Results

### Descriptive information on patients who had self-harmed

Across the study’s observation period (1st January 2019 to 31^st^ May 2021), 33,444 episodes of self-harm among 13,148 individuals were recorded; 59.5% (7,819) of individuals with a recorded self-harm episode were female, 3,320 (25.3%) were aged 10-17 years, 7,337 (55.8%) were aged 18-44 and 2,492 (19.0%) were aged 45 years and over. Around a fifth (2,893, 22.0%) of people who had self-harmed belonged to a non-White ethnic group. Among all 13,148 individuals who had self-harmed, IMD was missing for 22 (0.2%) of them and ethnicity was missing for 215 (1.6%). There were no missing data on age or sex.

### Overall temporal trends in monthly self-harm episode frequency counts

The frequency ratio of the number of incident self-harm episodes in April 2020 compared to February 2020 was 0.59 (95% CI 0.51 to 0.69) (Table 1a). Between August 2020 and May 2021 compared to the same months in 2019, the frequency ratio was 0.92 (CI 0.88 to 0.96) for incident episodes (Table 1a and Figure 1). In terms of total episodes (including multiple episodes by the same patients), the frequency ratios were 0.69 (CI 0.63 to 0.75) in April 2020 compared to February 2020 and 0.86 (CI 0.84 to 0.88) from August 2020 to May 2021 vs. the same months in 2019 (Table 1b and Figure 1).

**Table 1a:** Frequencies and ratio of frequencies in incident episodes between (i) April 2020 vs. February 2020 and (ii) August 2020 - May 2021 vs. August – Dec and Jan – May 2019.

|  | (i) April 2020 vs. February 2020 |  |  | (ii) August 2020 - May 2021 vs. August – Dec and Jan – May 2019 |  |  |
| --- | --- | --- | --- | --- | --- | --- |
|  | Frequency in April 2020 | Frequency in February 2020 | Ratio of frequencies (95% CI) | Frequency in pandemic study period | Frequency in 2019 comparison period | Ratio of frequencies (95% CI) |
| All (n) | 257 | 462 | 0.59 (0.51 to 0.69) | 4481 | 4877 | 0.92 (0.88 to 0.96) |
| Males(n) | 191 | 117 | 0.61 (0.49 to 0.77) | 1710 | 2039 | 0.84 (0.79 to 0.89) |
| Females | 158 | 271 | 0.58 (0.48 to 0.71) | 2770 | 2835 | 0.98 (0.97 to 1.03) |
| Ages 10-17 | 52 | 122 | 0.43 (0.31 to 0.59) | 1261 | 1181 | 1.07 (0.99 to 1.16) |
| Ages 18-44 | 156 | 258 | 0.60 (0.50 to 0.74) | 2439 | 2758 | 0.88 (0.84 to 0.93) |
| Ages 45+ | 67 | 82 | 0.82 (0.59 to 1.12) | 782 | 938 | 0.83 (0.76 to 0.92) |
| Deprivation quintile 1 (most deprived) | 153 | 240 | 0.64 (0.52 to 0.78) | 2067 | 2427 | 0.85 (0.80 to 0.90) |
| 2 | 55 | 86 | 0.64 (0.46 to 0.90) | 1072 | 1105 | 0.97 (0.89 to 1.06) |
| 3 | 25 | 55 | 0.45 (0.28 to 0.73) | 512 | 552 | 0.93 (0.82 to 1.05) |
| 4 | 25 | 41 | 0.61 (0.37 to 1.01) | 470 | 478 | 0.98 (0.87 to 1.12) |
| Deprivation quintile 5 (least deprived) | 16 | 40 | 0.40 (0.22 to 0.71) | 349 | 306 | 1.14 (0.98 to 1.32) |
| White | 208 | 346 | 0.60 (0.51 to 0.71) | 3404 | 3732 | 0.91 (0.87 to 0.96) |
| Black, Asian, mixed and other | 63 | 108 | 0.58 (0.43 to 0.80) | 1005 | 1067 | 0.94 (0.86 to 1.02) |

**Table 1b:**
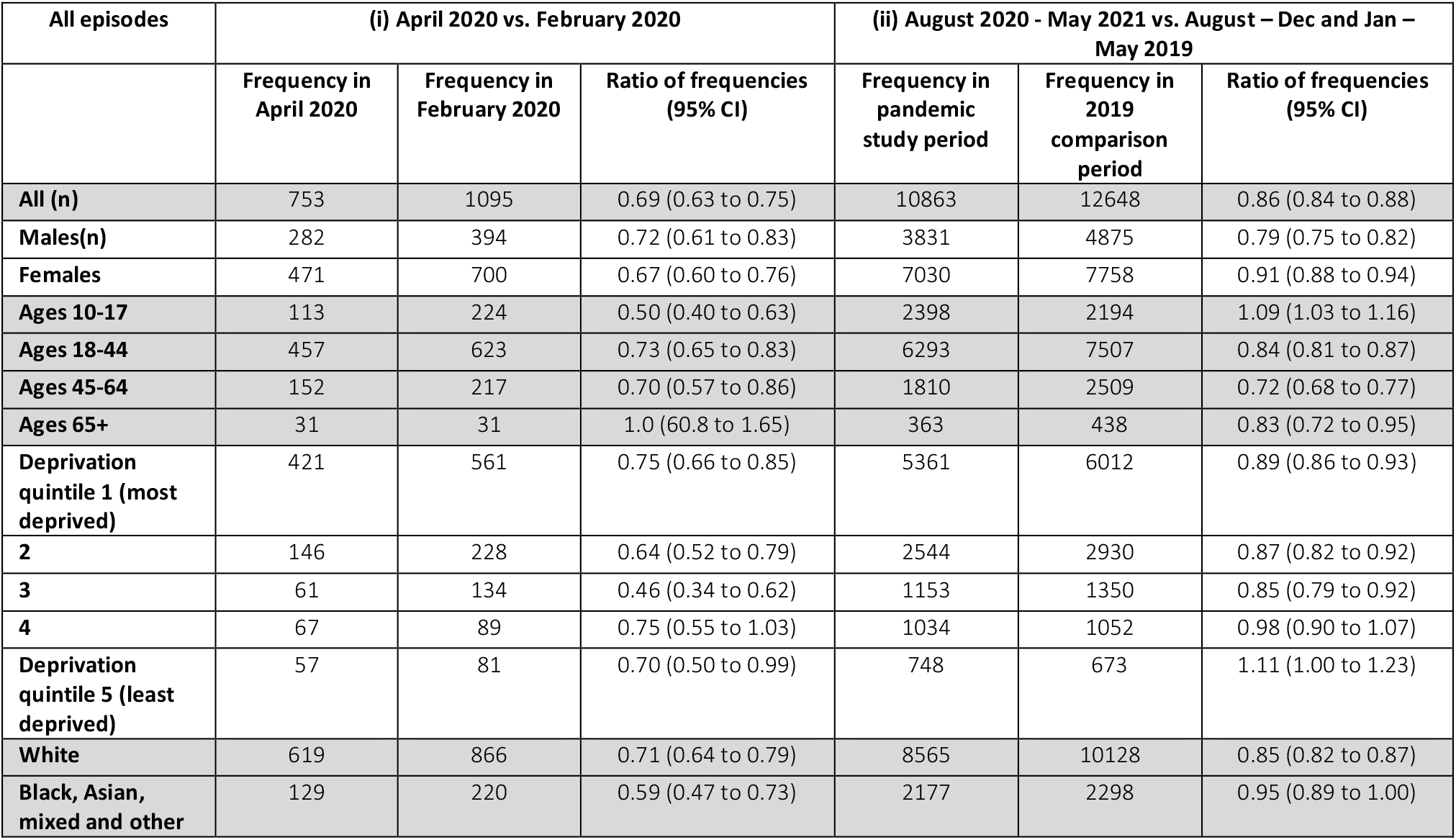
Frequencies and ratio of frequencies in all episodes between (i) April 2020 vs. February 2020 and (ii) August 2020 - May 2021 vs. August – Dec and Jan – May 2019.

**Figure 1:**
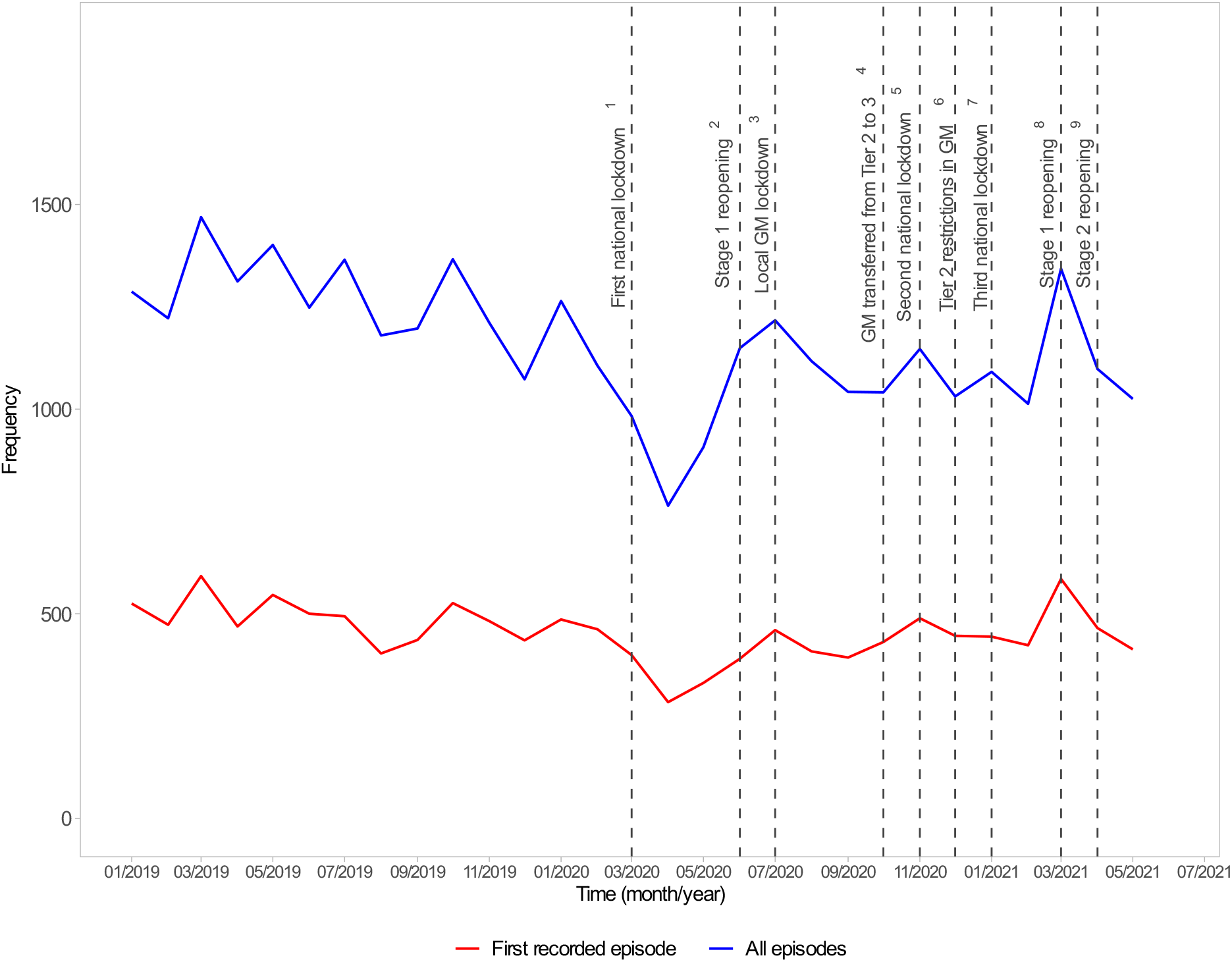
Temporal trends in monthly incident and all-episode self-harm frequency counts in relation to key national and regional COVID-19 restrictions, 1^st^ Jan. 2019 to 31^st^ May 2021 6: Tier 2 restrictions in GM equivalent to previous GM Tier 3

**Figure 2a & b:**
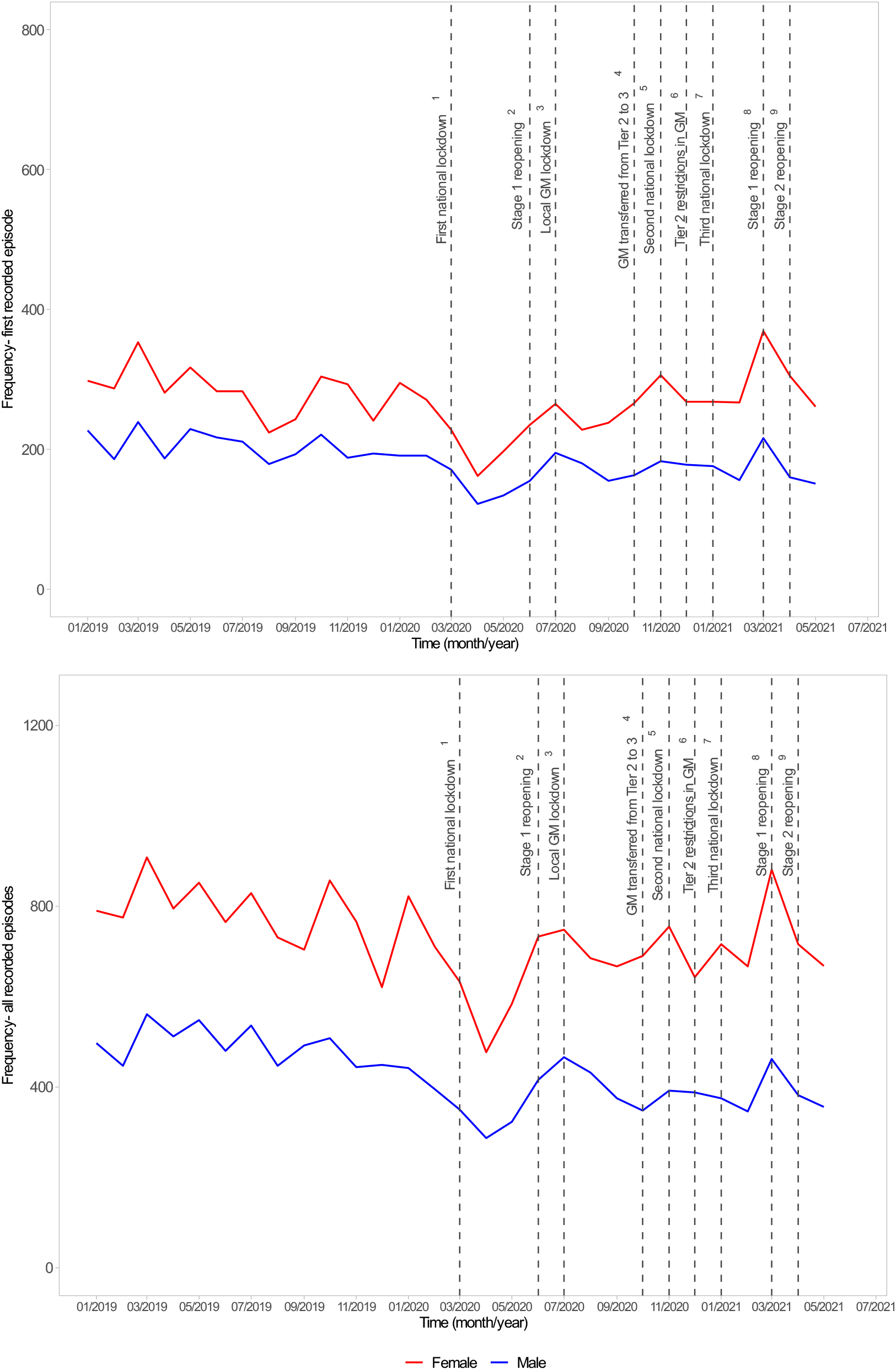
Sex-specific temporal trends in monthly incident and all-episode self-harm frequency counts with key national and regional COVID-19 restrictions, 1^st^ January 2019 to 31^st^ May 2021

**Figure 3a & b:**
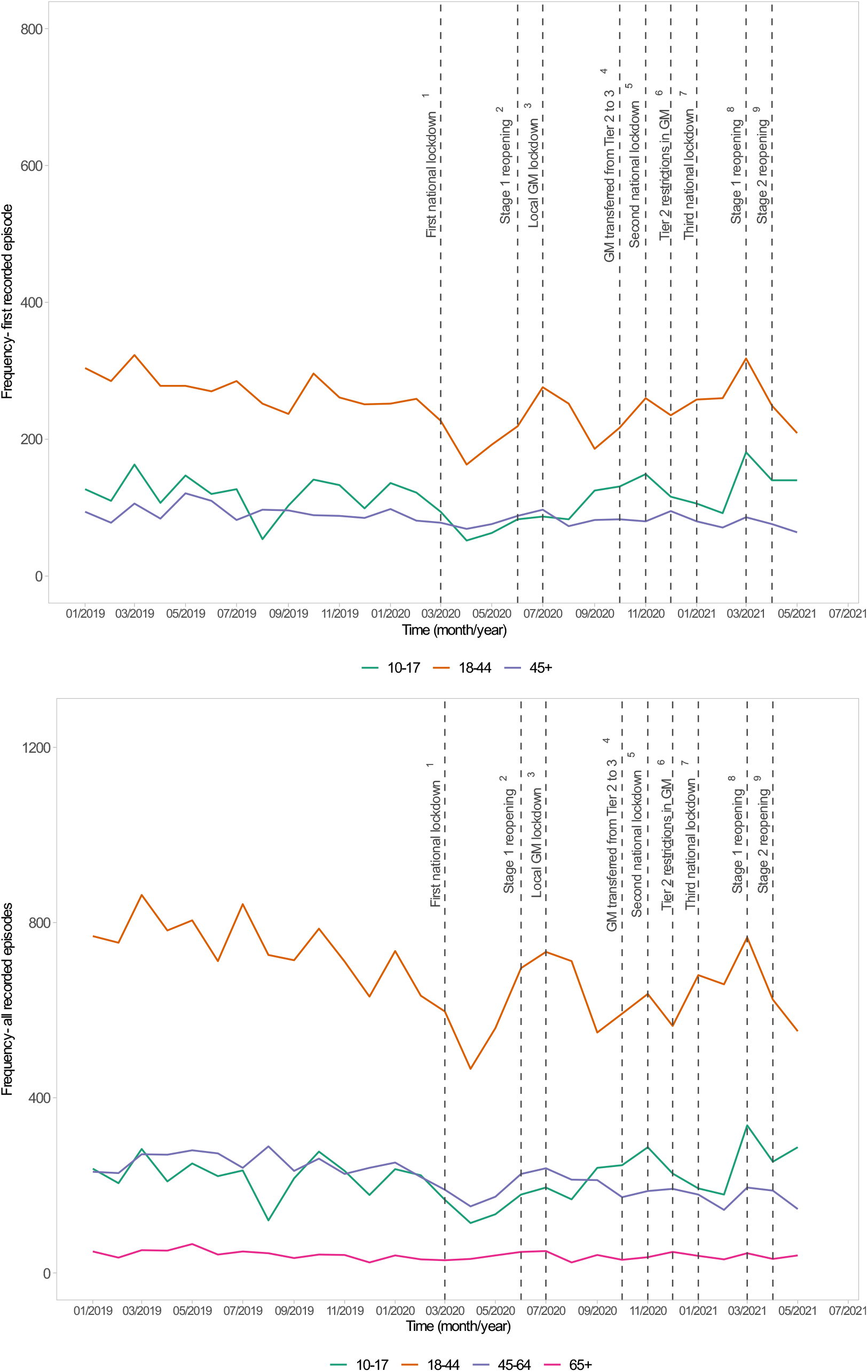
Age-specific temporal trends in monthly incident and all-episode self-harm frequency counts with key national and regional COVID-19 restrictions, 1^st^ January 2019 to 31^st^ May 2021

### Temporal trends in monthly self-harm episode frequency counts by sex, age group, deprivation quintile and ethnicity

In April 2020 the reductions in incident self-harm were similar among women and men (Table 1a). Monthly frequencies of first self-harm episodes increased among both groups between August 2020 and May 2021. Among women, episode counts were similar to 2019 (frequency ratio 0.98 (0.97 to 1.03) while among men, a reduction remained (Table 1a). In terms of all episodes of self-harm, a larger reduction was also observed among men during August 2020 to May 2021 (frequency ratio 0.79 (CI 0.75 to 0.82) vs. 0.91 (CI 0.88 to 0.94) among women.

In April 2020, the frequency ratios in incident and all episode self-harm counts among young people aged 10-17 years were 0.43 (CI 0.31 to 0.59) and 0.50 (0.40 to 0.63) respectively (Table 1a and b), lower than for other age groups. Between August 2020 and May 2021, there was an increase in incident (1.07, CI 0.99 to 1.16) and all self-harm episodes (1.09, CI 1.03 to 1.16) among patients aged 10-17. Among people living in more deprived neighbourhoods, we observed reductions in both incident and all episode self-harm counts in the August 2020 to May 2021 period. For example, in the most deprived quintile, the frequency ratio of first self-harm episodes was 0.85 (CI 0.80 to 0.90) compared to 1.14 (CI 0.98 to 1.32) in the least deprived quintile (Figures 4a and b and Table 1a). The reductions in all recorded episodes in April 2020 were observed in the two broad White and non-White ethnicity categories (Figures 5a and 5b and Table 1a and b) The longer-term reduction was larger in the White group (frequency ratio 0.85 (0.82 to 0.87) vs. 0.95 (0.89 to 1.00) in the non-White group.

**Figure 4a & b:**
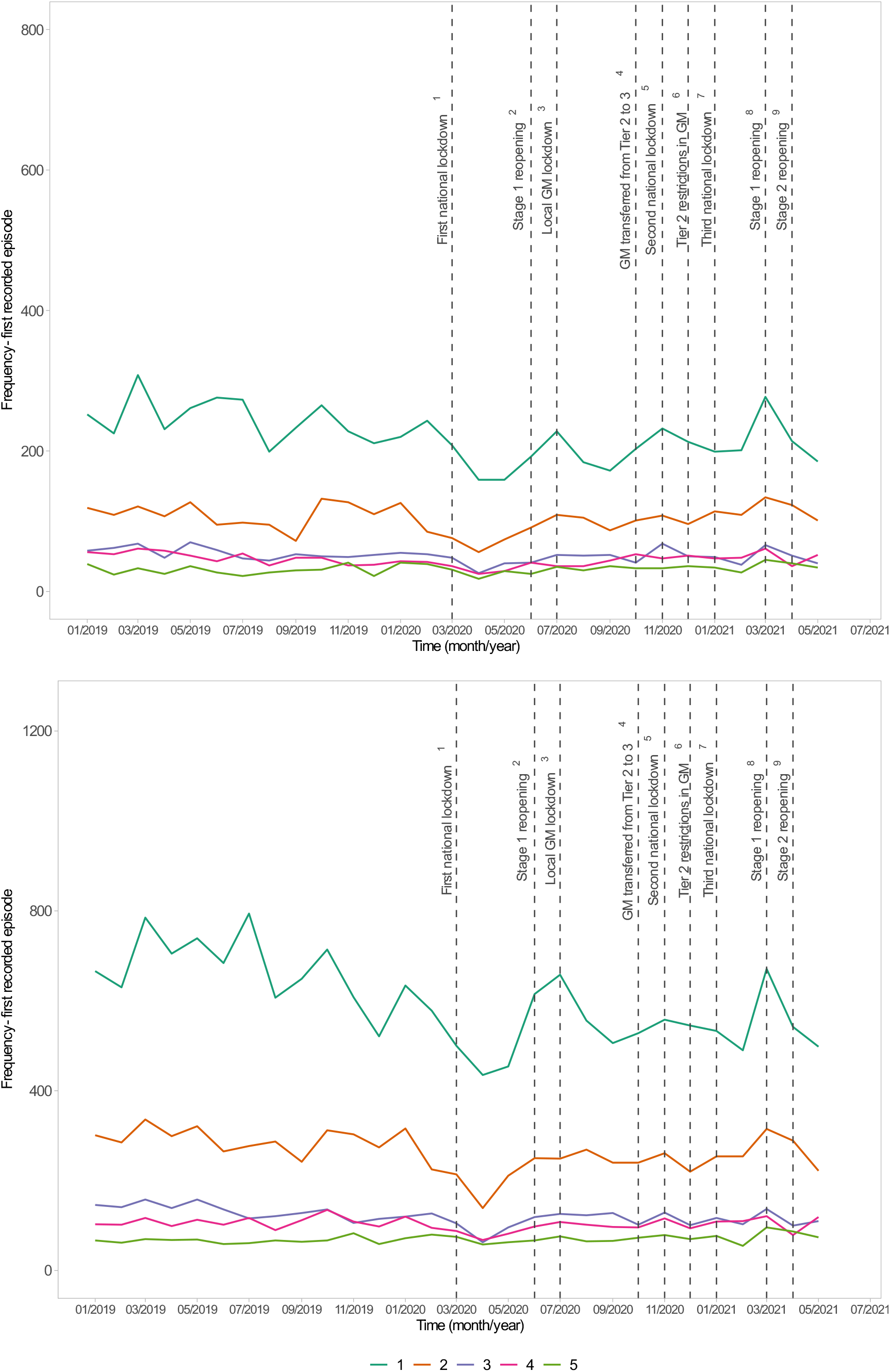
Deprivation quintile-specific temporal trends in monthly incident and all-episode self-harm frequency counts with key national and regional COVID-19 restrictions, 1^st^ January 2019 to 31^st^ May 2021

**Figure 5a & b:**
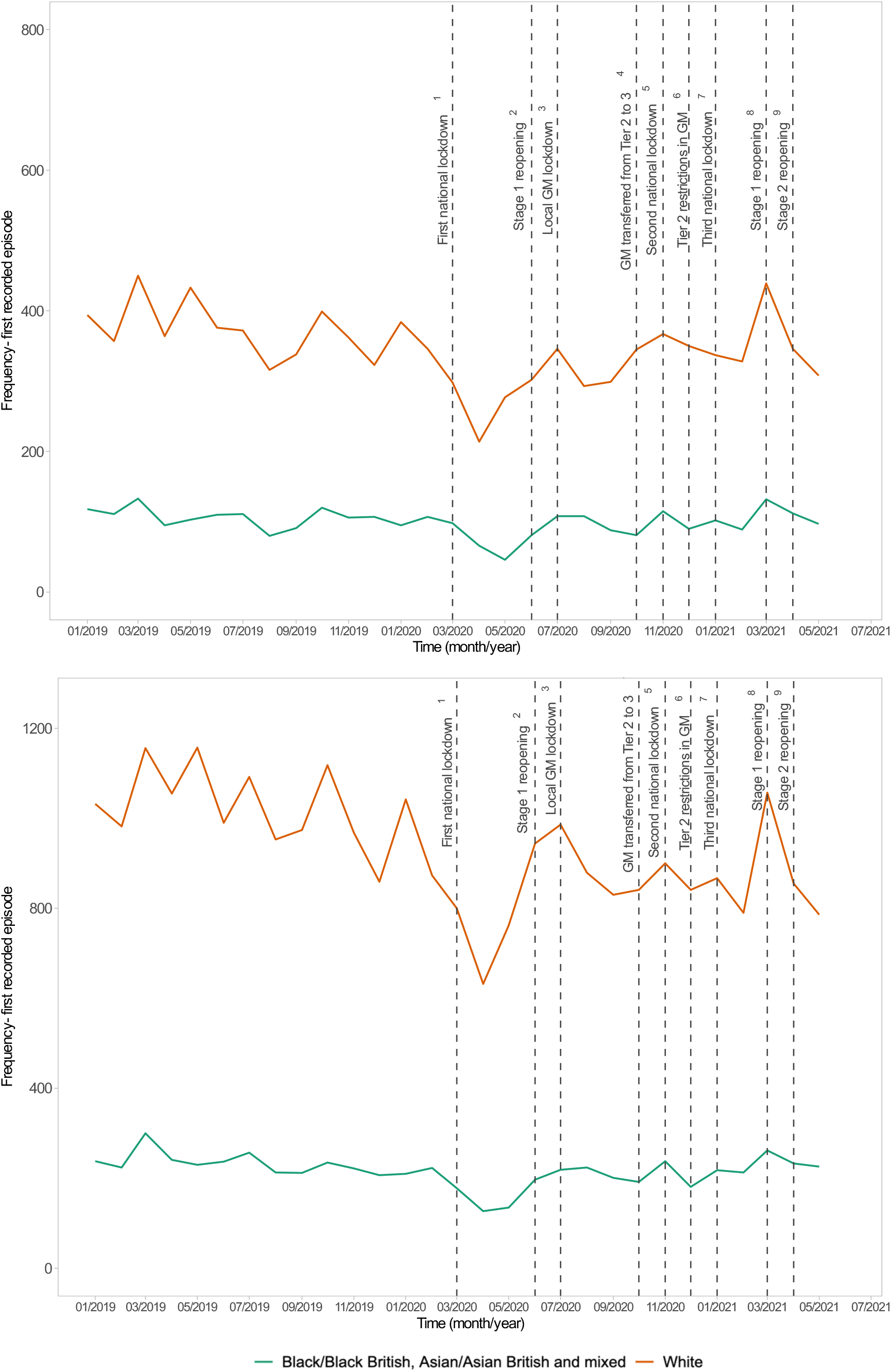
Ethnic group-specific temporal trends in monthly incident and all-episode self-harm frequency counts, with key national and regional COVID-19 restrictions, 1^st^ January 2019 to 31^st^ May 2021

### Overall self-harm episode frequency counts by phases of national and regional restrictions

The frequency ratios were lowest during months when the region was experiencing the first national lockdown: incident episodes 0.62 (CI 0.58 to 0.67) and all episodes 0.63 (CI 0.60 to 0.66). Frequency ratios attenuated in November and December 2020 so that incident and all episode counts were in line with those from 2019 (Table 2). In the subsequent phase representing a period of national lockdown (January to February 2021) frequencies of incident and all recorded self-harm were lower than the same months in 2019, though the falls in frequency were smaller than during the first lockdown. During the period including the phased easing of restrictions (March to May 2021), numbers of incident self-harm episodes were in line with 2019 (0.98 (CI 0.90 to 1.07), though the total episode count remained lower: 0.87 (0.83 to 0.92).

**Table 2:** Ratio of frequencies in incident and all episodes by periods^1^ of national and regional Covid-19 restrictions.

| Time period | Incident episodes |  |  | All episodes |  |  |
| --- | --- | --- | --- | --- | --- | --- |
|  | Frequency in pandemic study period | Frequency in 2019 | Ratio of frequencies (95% CI) | Frequency in pandemic study period | Frequency in 2019 | Ratio of frequencies (95% CI) |
| **1<sup>st</sup> March to 31<sup>st</sup> May 2020:** first national lockdown | 999 | 1,605 | 0.62 (0.58 to 0.67) | 2,619 | 4,168 | 0.63 (0.60 to 0.66) |
| **1<sup>st</sup> June to 30<sup>th</sup> July 2020:** stage 1 of reopening | 845 | 995 | 0.85 (0.77 to 0.93) | 2,348 | 2,609 | 0.90 (0.85 to 0.95) |
| **1<sup>st</sup> August to 31<sup>st</sup> October 2020:** GM regional restrictions and schools reopening<sup>2</sup> | 1,227 | 1,361 | 0.90 (0.83 to 0.97) | 3,176 | 3,724 | 0.85 (0.81 to 0.89) |
| **1<sup>st</sup> November to 31<sup>st</sup> December 2020:** second national lockdown and regional restrictions<sup>3</sup> | 937 | 916 | 1.02 (0.93 to 1.12) | 2,175 | 2,271 | 0.96 (0.90 to 1.02) |
| **1<sup>st</sup> January to 29<sup>th</sup> February 2021:** third national lockdown<sup>4</sup> | 863 | 995 | 0.87 (0.79 to 0.95) | 2,069 | 2,485 | 0.83 (0.79 to 0.88) |
| **1<sup>st</sup> March to 31<sup>st</sup> May:** phased easing of restrictions including schools and colleges reopening<sup>5</sup> | 1,042 | 1,059 | 0.98 (0.90 to 1.07) | 2,419 | 2,775 | 0.87 (0.83 to 0.92) |

<sup>1</sup> Phases are represented by the main calendar months they occurred in as counts were not available by specific dates.
<sup>2</sup> Full reopening of schools (September 2020), introduction of regional 'tiered' restrictions: 14th October 2020 (GM: Tier 2); 23<sup>rd</sup> October 2020: GM transferred from Tier 2 to 3
<sup>3</sup> National lockdown: 5<sup>th</sup> November 2020; Tier 2 (equivalent to previous Tier 3) restrictions in GM: 2<sup>nd</sup> December 2020.
<sup>4</sup> Third national lockdown: 6<sup>th</sup> January 2021
<sup>5</sup> Stage 1 of phased easing of restrictions (including all schools and colleges reopening): 8<sup>th</sup> March 2021; Stage 2 of phased easing of restrictions: 12th April 2021; Stage 3 of phased easing of restrictions: 17th May 2021.

## Discussion

### Main findings

Following an initial marked reduction in overall frequency of primary care recorded self-harm in April 2020, a sustained reduction was observed to the end of May 2021. These longer-term reductions were largest among men and people living in the most deprived neighbourhoods, though there was an increase in recorded self-harm among adolescents aged 10-17 years. The greatest overall reductions were observed during months when the Greater Manchester conurbation was experiencing the first national lockdown. In the last two months of 2020, the period covering the second national lockdown and regional restrictions, overall numbers of incident self-harm episodes were in line with 2019, though further reductions followed in January and February 2021.

### Interpretation and comparison with existing evidence

The reductions in monthly incident and all-episode self-harm frequency during April 2020 were broadly similar to those found in a study of general practices across the UK that was conducted in the Clinical Practice Research Datalink (CPRD). ^9^ In that study, women and patients registered at practices located in the most deprived quintile had greater reductions, as we also found. However, while that CPRD-based study reported that monthly episode counts were broadly in line with expected levels by July 2020, ^9^ our findings show evidence of sustained reductions in men and people living in more deprived areas. Many UK residents whose mental health deteriorated after the pandemic began reported experiencing an improvement by October 2020. ^11^ However, people with current or previous mental health problems and those living in deprived neighbourhoods were overrepresented among those individuals whose mental health did not improve or worsened.^11^ This suggests that the prolonged reductions in the volume of people seeking help for self-harm that our study in Greater Manchester has revealed did not simply reflect a reduced need for services.

We found that the second and third national lockdowns, as well as the regional restrictions experienced by people in Greater Manchester, had a less profound impact on the numbers of people treated for self-harm than the first national lockdown beginning in March 2020. This may reflect greater willingness to use health services as confidence in health services to cope with demand increased. During the period November to December 2020, when the second national lockdown and regional restrictions were in place, the numbers of primary care recorded self-harm were in line with those in 2019, suggesting attitudes to help seeking were at pre-pandemic levels. Attitudes are also likely to be associated with rates of COVID-19 hospitalisations, and may have been influenced by the higher hospitalisation rates in the first weeks of 2021. ^23^ These findings may also reflect increased clinical need for self-harm during the Winter 2020 period.

The reductions in episode frequency counts that we have observed may indicate missed opportunities for treating people who self-harmed but did not present to health services, as well as a potential real fall in incidence in the population. The more prolonged reduction in recorded self-harm that we observed could reflect people seeking help from alternative sources such as online networks, social media, third sector helplines or engagement in alternative coping mechanisms. ^2^ While comparisons with pre-pandemic periods are challenging, one study suggested a lower proportion of adults in the UK who had self-harmed during March and April 2020 sought help from a medical professional and a higher proportion reported speaking about their mental health with friends or family. ^24^ However, evidence from France showed that, while overall number of hospitalisations for self-harm decreased in the early months of the COVID-19 pandemic, severity of self-harm was higher. ^25^ This suggests some affected individuals may not have received any support. The provision of good quality psychosocial assessment by a mental health specialist is recommended following any self-harm episode. Reduced frequency in presentation to services for self-harm suggests that people have been less likely to receive this care. ^20^ Likelihood of referral to mental health services from primary care among people who had self-harmed was found to be lower than in previous years during the first three months of the pandemic, while likelihood of being prescribed with psychotropic medication by a GP or practice nurse was slightly higher. ^26^ Considering the potential increased demand for psychosocial interventions for self-harm in some groups during the pandemic, timely access to appropriate services should be prioritised.

In our study of primary care electronic health records, the reduction in incident episodes of self-harm observed among women in April 2020 appears to have returned to 2019 levels. Self-harm occurs more commonly in women than men ^27^ and the higher level of clinical need and complexity associated with deprivation, alongside the evidently greater detrimental effect of the pandemic on women’s mental health, ^10^ may explain this finding. Evidence also suggests that women have experienced worse mental health during the pandemic than men. ^10,28^ However, the potential treatment gap among men is also a major concern, particularly given the much higher risk of suicide among men who have harmed themselves. ^29^ We also found that self-harm frequency counts among 10-17 year olds were higher in the period August 2020 to May 2021 than the same months in 2019. While the pandemic is likely to have affected adolescents’ mental health, ^30^ it is not known to what extent the increase we found reflects the existing trajectory of worsening mental health among adolescents prior to the pandemic.^31^ In any case, self-harm is the strongest risk factor for suicide in adolescents and young people ^32^ and early intervention for young people who have self-harmed is recommended. ^33^

Continued monitoring of frequency counts for recorded self-harm episodes is important in understanding how perceptions of the accessibility of general practice and hospital emergency departments has fluctuated through the course of the pandemic, as well as providing insight into gaps in help-seeking that have arisen and potential increases in clinical need. Examining presentations to emergency departments, and including follow-up time to quantify future risks of dying by suicide and by other external causes following self-harm, will help in answering important questions about whether COVID-19 has affected the severity of patients who do present, for example in terms of future suicide risk. This will also help in understanding the potential impact of missed opportunities for treatment resulting from non-presentation to services.

## Strengths and limitations

Utilisation of the GMCR offered a number of unique advantages. The near ‘real-time’ data availability enabled us to access more up-to-date evidence than is available in other data sources. The data source enabled us to study 2.8 million residents in Greater Manchester registered with a GP, meaning that our findings are representative of patients across this large and socially diverse conurbation. Furthermore, Greater Manchester is of particular interest due to the heightened regional and ‘tiered’ COVID-19 containment restrictions that the conurbation experienced from July 2020.

We could not examine monthly patterns in frequency counts prior to 2019 due to unavailability of data for earlier years in the GMCR. However, we were able to use historic patient records to check for previous episodes of self-harm, enabling us to identify incident self-harm in the study population. Our examination of self-harm episodes by phases of restrictions were represented by the main calendar months they occurred in as counts were not available by specific dates. Our findings may not be representative of other areas of the UK, particularly those that did not experience similar regional COVID-19 containment restrictions. We were not able to examine Black/Black British, Asian/Asian British and other ethnic groups separately due to low monthly episode counts (to prevent risk of disclosure we could not present cell counts lower than 10).

## Conclusions

The COVID-19 pandemic and associated national and lockdowns and regional restrictions persisting into the spring of 2021 have had a prolonged impact on help seeking for self-harm. Reductions among those living in more deprived neighbourhoods is evidence that deepening of pre-pandemic health inequalities is persisting. The larger reduction in recorded self-harm among men suggests a potential treatment gap. During the ten months leading to May 2021, adolescents aged 10-17 years were more likely to have an episode of self-harm recorded in primary care than in the same months in 2019 suggesting the clinical need among this group has increased. The trends we observed suggest the COVID-19 pandemic has implications for clinicians’ ability to assess the needs and risks of individuals. Some patients may have experienced untreated deterioration in their mental health up to over a year after the first wave of the pandemic. There are also important implications for potential demand experienced by primary care and mental health services.

## Data Availability

The clinical codes used in this study are available online at https://github.com/rw251/gm-idcr/tree/master/shared/clinical-code-sets/patient/selfharm-episodes/1. The codes are also available from the corresponding author on request. Access to data are only available upon approval by the GMCR Research Governance Group.

## Acknowledgements

The authors recognise the Greater Manchester Care Record (a partnership of Greater Manchester Health and Social Care Partnership, Health Innovation Manchester and Graphnet Health, on behalf of Greater Manchester localities) in the provision of data required to undertake this work. We thank Stephen Barlow, Elizabeth Monaghan, Fiona Naylor, and Jonathan Smith (members of the Centre for Mental Health and Safety patient and public involvement and engagement group of the NIHR Greater Manchester Patient Safety Translational Research Centre) for their advisory roles in the study. This work uses data provided by patients and collected by the NHS as part of their care and support.

## Author contributions

All authors conceptualised the study and contributed to its design. LB, GT and SS managed the data, did statistical analysis and verified the data. SS drafted the manuscript. All authors critically reviewed the manuscript and approved the final version.

